# Hearing Progression of Children with Enlarged Vestibular Aqueduct (EVA) According to *SLC26A4* Genetic Mutation Status

**DOI:** 10.1101/2023.11.29.23299192

**Authors:** Madeline Hocking Osborne, Margaret Koeritzer, Hannah Herd, Melissa Jane Polonenko

## Abstract

Enlarged vestibular aqueduct (EVA) is a malformation associated with sensorineural hearing loss in children. Mutations in the *SLC26A4* gene are also associated with EVA and may contribute to more severe hearing losses that progress, but the timescale of this progression remains unknown. Given that children undergo significant speech and language development, a better understanding of the timing and extent of hearing loss is needed for individuals who have EVA. Through a retrospective review of records in a local database, we aimed to 1) estimate the prevalence of a *SLC26A4* mutation in this pediatric EVA cohort; and 2) compare hearing threshold severity and timing of progression across children who have EVA and different *SLC26A4* statuses. Of 62 children with EVA, 39 had available genetic results and of these, 15 had at least one mutation of the *SLC26A4* gene (38%). The children with a *SLC26A4* mutation had a more severe onset of hearing loss and a more rapid progression of hearing loss over time, especially for higher frequencies, than children without a mutation in this gene. This longitudinal information may facilitate prognostic counseling to pediatric patients and better inform the audiology follow-up appointments for these families.

## Introduction

There are several causes of progressive hearing loss in children. One such cause in an enlarged bony canal and membranous labyrinth that leads out of the vestibule of the inner ear and into the temporal bone, called enlarged vestibular aqueduct (EVA). EVA is the most common inner ear malformation in children and accounts for 15 to 32% of all pediatric causes of sensorineural hearing loss (Hodge et al. 2021). Hearing loss associated with EVA tends to be bilateral and progressive or fluctuating, but the precise way the malformation causes hearing progression remains unknown.

One factor that might contribute to the time course of hearing loss associated with EVA is the association with mutations of the *SLC26A4* gene. This gene gives rise to both non-syndromic and syndromic EVA, often associated with Pendred syndrome. Approximately 50% of all individuals with EVA who have hearing loss also have a mutation in the *SLC26A4* gene (Rose et al. 2017). A mutation in the *SLC26A4* gene may also contribute to hearing losses that are more likely to be progressive and severe (King et al. 2010).

Previous studies have shown that individuals with EVA who have mutations in the *SLC26A4* gene tend to have more severe hearing loss compared to those without mutations. In a prospective cohort survey of 83 people with EVA, most of whom were adults, the severity of hearing loss strongly correlated with the number of mutant alleles in the *SLC26A4* genotype (King et al., 2010). Specifically, individuals with EVA with zero or one mutant allele had less severe hearing loss, while those with two mutant alleles had more severe hearing loss (King et al 2010). Besides the number of mutations, hearing severity was not associated with any other factors, including goiters from Pendred syndrome or presence of other cochlear malformations.

A follow-up study with 127 individuals by the same group showed that the number of mutant alleles of *SLC26A4* significantly correlated with hearing loss progression in addition to the severity of hearing loss. Ears with EVA and zero or one mutant alleles had mean 4-frequency pure tone air conduction averages (PTA; 500–4000 Hz) of 63- and 53-dB HL, respectively, compared to 88 dB HL for those with two mutant alleles (Rose et al. 2017). Furthermore, the prevalence of having a cochlear implant significantly increased with number of mutations: 12%, 13%, and 38% for zero, one, and two mutant alleles, respectively (Rose et al. 2017). The higher prevalence of cochlear implantation in individuals with EVA and two mutant alleles of *SLC26A4* likely relates to the greater severity of hearing loss found in these same individuals. This study concluded that individuals with EVA and zero or one mutant allele of *SLC26A4* have less severe hearing loss, less progression of hearing loss, and a lower prevalence of cochlear implantation, compared to individuals with two mutant alleles of *SLC26A4*. As a result, *SLC26A4* mutation status appeared to be predictive of both severity of hearing loss and amplification options.

While these studies largely highlight the increased severity and prevalence of hearing loss progression experienced by adults with EVA, there are two main gaps in our knowledge that are important for clinical management of EVA. First, these studies primarily include adults, but EVA is a congenital malformation. More data is needed to examine the relationship between mutations in the *SLC26A4* gene, EVA, and the progression of hearing loss in a pediatric-specific population, which would better inform the management strategies for children with EVA. Second, in each of these studies, hearing loss progression was only categorized as a binary outcome of absent or present. The exact time course of this hearing progression remains unknown. As such, there is little information available to better guide counseling for pediatric patients about the rate at which the hearing loss may progress. Months versus years versus decades has significantly different implications for the pediatric population, as they are undergoing critical periods of hearing development, as well as spoken speech and language acquisition. Furthermore, a largely unknown rate of hearing progression can be a very confusing diagnosis for families. Monitoring the hearing loss should be appropriate to detect changes and make appropriate adjustments to amplification, but also not become burdensome to the family.

Because progressive hearing loss is commonly observed in people with EVA, close audiologic monitoring is warranted. Current clinical management of EVA is largely reactive, meaning that clinicians often wait until a “crisis” or drop in hearing occurs. Being proactive is challenging because outcomes are variable, and we know that progression *may* occur but not *when* or *by how much*. Thus, there is an unmet need to improve our understanding of EVA by exploring potential clinical prognostic factors for hearing loss severity and progression (Saeed et al. 2022). Understanding the implications of their hearing loss is crucially important as they progress through social and communication developmental periods. If we know more about the differences in hearing loss among children who have EVA both with and without an *SLC26A4* genetic mutation, then we will be better equipped to provide prognostic counseling to affected children and their families.

Given the unknown rate of hearing progression and its potential clinical value, this study involved a retrospective review of records of all children with EVA in our local clinical database. <u>The study aims were to:</u> 1) estimate the prevalence of an *SLC26A4* mutation, 2) compare severity of hearing thresholds across children who have EVA and different *SLC26A4* status and 3) compare the rate of change in hearing thresholds across children who have EVA and different *SLC26A4* status. <u>We hypothesized</u> that children with EVA who possess the *SLC26A4* gene mutation will have an earlier onset of a more severe hearing loss that progresses more rapidly compared to children with EVA who do not possess the *SLC26A4* gene mutation. *Understanding these audiological markers will be of significant clinical value to families by better informing counseling, follow-up recommendations, and understanding potential risks during key speech and language developmental periods*.

## Methods

We retrospectively identified children with EVA from the ∼500 records currently in the “*Database of Infants and Children Seen at the Lions Children’s Hearing Center at the University of Minnesota”.* Institutional Review Board approval was obtained from the University of Minnesota and Fairview Health Systems Research (Study 16972). De-identified data were reviewed from the database’s creation in 2012 until August 31, 2022. Inclusion criteria were ages up to 18 years, a diagnosis of EVA as confirmed by imaging (Computerized Tomography or Magnetic Resonance Imaging), and hearing thresholds from at least two visits to allow for a longitudinal analysis. With this criteria, 62 children (12.4%) were included for further analysis.

Audiometric information from each of the 62 patients was collected. This included: 1) case history, 2) newborn hearing screening results, 3) which ear(s) had EVA, 4) the type of test conducted in each visit (e.g., auditory brainstem response, conditioned play audiometry, or visual reinforcement audiometry), 5) air and bone conduction hearing thresholds in the left and right ear(s) at 500, 1000, 2000, and 4000 Hz, when available, 6) *SLC26A4* genetic testing results, 7) cochlear implantation status, 8) date of birth, and 9) date at each visit.

The *SLC26A4* genetic testing results were sorted into three categories: a) Mutation (known to be related to hearing loss), b) No Mutation, and c) Unknown (genetic testing not conducted or available for the *SLC26A4* gene). The unknown group was included for comparison because lacking access to genetic results reflects the reality of most clinics. The de-identified data was organized, curated, and analyzed using Microsoft Excel and RStudio (RStudio Team 2020).

The Shapiro-Wilks test was conducted to determine if variables were normally distributed and whether to perform parametric or nonparametric analyses. The extent of change in hearing threshold severity over time across the three groups was analyzed using linear mixed effects models to account for the variability of each child and repeated data. A two-way proportions test was used to compare percentage of children who had severe hearing loss at initial versus last available visits, and Kruskal-Wallis tests were used to analyze hearing severities between groups for the first and last visits, and when appropriate, Wilcoxon signed-rank tests were done as post-hoc tests. The following R packages were used: lmerTest (v3.1-3; Kuznetsova et al 2020), lme4 (v1.1-35.1; Bates et al 2023), PerformanceAnalytics (v2.0.4; Peterson et al 2020), buildmer (v2.11; Voeten 2023).

## Results

### Variability among children with EVA according to genetic status

The first aim of this study was to estimate the prevalence of a deleterious *SLC26A4* mutation in our local population of children with EVA. Of the 62 children with EVA, 15 had at least one confirmed mutation in the *SLC26A4* gene (24%), 24 were confirmed to have no mutation in this gene (39%), and 23 had not been genetically tested for this gene and were thus classified as “Unknown” (37%). Of the children that were genetically tested, 38% (15/39) had at least one *SLC26A4* mutation. See **Table 1** for details.

**Table 1.** Summary of Selected Clinical Population of Children with EVA by SLC26A4 Status.

| **Group** | **No. Children (% total, n=62)** | **Age at first visit Median (range) years** | **Age at last visit Median (range) years** | **Time between visits Median (range) years** | **No. children (%) with Cochlear Implant(s)** |
| --- | --- | --- | --- | --- | --- |
| **Mutation** | 15 (24%) | 2.8 (0–9) | 5.4 (1–9) | 2.8 (0–6) | 13 (87%) |
| **No Mutation** | 24 (39%) | 3.6 (0–16) | 6.8 (0–18) | 2.9 (0–10) | 8 (33%) |
| **Unknown (no genetic testing)** | 23 (37%) | 2.5 (0–14) | 4.9 (1–15) | 2.4 (0–11) | 16 (70%) |

The children’s ages at first and last visits are also displayed in **Table 1**. For this cohort, the age at first visit for many of the children depended on when the hearing loss was first diagnosed. The median ages for the Mutation, No Mutation, and Unknown groups were 2.8, 3.6, and 2.5 years respectively for the first visit, and 5.4, 6.8, and 4.8 years respectively for the last visit. The ages for each group were not significantly different from each other (Kruskal-Wallis, first visit *χ^2^*(2)=0.6, *p*=0.742; last visit *χ^2^*(2)=1.7, *p*=0.433). Consequently, the durations between first and last visits were also not statistically different for the Mutation, No Mutation, and Unknown groups: 3.7, 4.1, and 4.4 years respectively (Kruskal-Wallis, *χ*^2^(2)=1.9, *p*=0.383).

The type of amplification used by the children reflects the severity of their hearing loss.

Children with a more severe hearing loss are more likely to receive a cochlear implant rather than continue with using a hearing aid. The proportion of children with cochlear implants in the Mutation group was higher (86.7%) than the No Mutation (33.3%) or Unknown (69.6%) groups (Proportions test, *χ^2^*(2)=12.4, *p*=0.002), although only the corrected post-hoc comparison between the Mutation and No Mutation proportions of children with CIs was significant (Bonferroni corrected p=0.010; comparisons to Unknown group, corrected p>0.08).

Furthermore, children with EVA and a *SLC26A4* mutation were 2.6 times more likely to obtain a CI than children without a mutation, although this relative risk ratio was not statistically significant using a conservative assessment, given the smaller cohort size (Fisher test; *p*=0.107). Alternatively, a child with EVA and a CI was 12.1 times more likely to be from the Mutation group than No Mutation group (Odds Ratio p=0.002).

### Hearing loss is more severe for children with a mutation

The second aim of this study was to compare hearing thresholds across children with EVA based on their *SLC26A4* status. **Figure 1A** shows histograms highlighting the number of children in each group with specific ranges of hearing thresholds, quantified by pure tone average (PTA), at their first and last recorded visits. At the first visit, both the No Mutation and Unknown groups had PTAs that spanned a wide range of severities, from 0 dB HL (normal hearing) to 120 dB HL (profound hearing loss). In contrast, the Mutation group had a much narrower distribution of hearing thresholds, which concentrated around severe hearing loss (> 70 dB HL). Therefore, children with a *SLC26A4* mutation tended to have poorer hearing at their first visit than those without a mutation, however this difference was not statistically significant (medians of 74.4 dB HL, 53.8 dB HL and 70.3 dB HL, respectively; Kruskal-Wallis test: *χ^2^*(2)=3.89, *p*=0.142). At the last visit, the No Mutation group remained widely distributed across the range of severities, whereas the Mutation and Unknown groups showed a significant shift in the distribution of PTAs towards higher PTAs, or worse hearing, relative to the first visit distribution, indicating more children had severe/profound hearing loss. The last visit median PTAs for the Mutation, No Mutation and Unknown mutation groups were 95.8 dB HL, 52.9 dB HL, and 100.6 dB HL, respectively (Kruskal-Wallis test: *χ*^2^(2)=7.15, *p*=0.028; Bonferroni-corrected post-hoc Wilcoxon signed rank sum test between Mutation and Unknown p>0.05, and p<0.04 for comparisons to the No mutation group). Although the Unknown group showed shifts in PTAs and more severe losses at the second visit, the group tended to remain more variable (spread) than the Mutation group. However, the No Mutation group was clearly the most variable, making it difficult to predict the severity of their hearing loss and inform counseling decisions based on their *SLC26A4* status.

**Figure 1.**
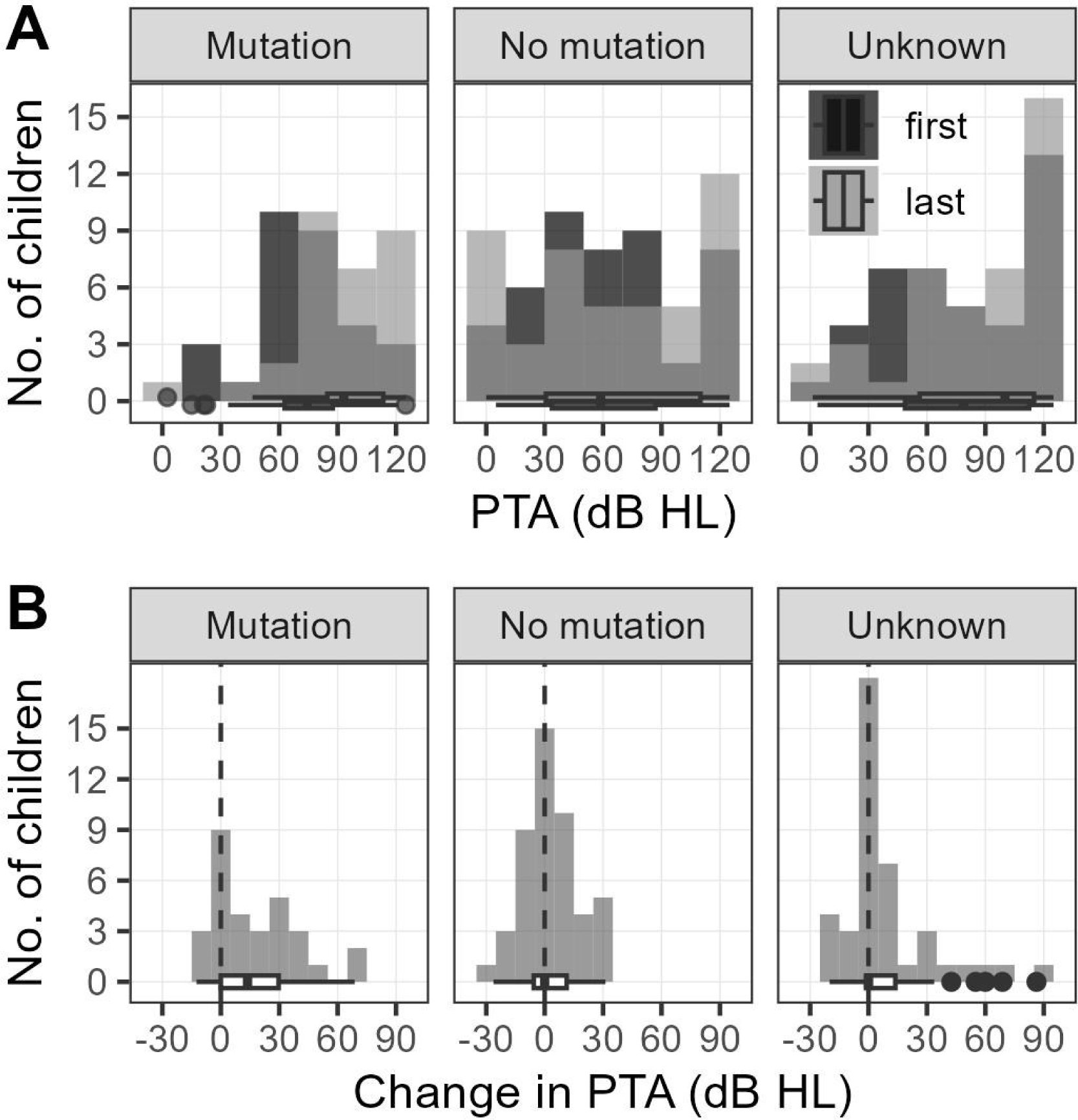
Group distributions of A) pure tone average (PTA) thresholds at first and last visit, and B) change in PTA thresholds from first to last visits.

The shift in hearing loss severity was further explored in **Figure 1B**, which shows histograms and boxplots of the change in PTA from the first to last visit for the three groups. The dashed vertical line represents no change from first to last visit (i.e., a 0 dB HL change in PTA). For the Mutation group, most of the data is spread to the right of the dashed line, indicating that there was an overall worsening of the PTA for most children in this group. The median change in PTA for the Mutation group was 13.7 dB HL (from first to last visit). By comparison, both the No Mutation and Unknown groups were centered around the dashed line and distributed evenly on both sides. The box plot for both of these groups shows their medians to be around 0 dB HL (with outliers for the Unknown group), indicating little to no change overall in PTA from the first to last visit in the No Mutation and Unknown mutation groups (each individual child may still have experienced a shift in PTA in either direction). In fact, our post-hoc testing revealed that there was a significant difference in the change in hearing threshold between first and last visit for the Mutation group when compared to the No Mutation group (Kruskal-Wallis test: *χ^2^*(2)=9.46, p=0.009: Wilcoxon signed rank sum post-hoc test: *W*(1)=1001, corrected *p*=0.006).

In addition, we also analyzed the shift in severity of hearing loss by calculating the percentage of ears with severe hearing loss for each visit and group (**Figure 2**). The Mutation group had a marginally higher percentage of ears with severe hearing loss at the first visit (63.3%) compared to the No Mutation and Unknown groups (40.4% and 53.6%, respectively; proportion test: *χ^2^*(2)=4.04, *p*=0.13), but a significantly higher proportion of 86.7% with severe hearing loss by the last available visit compared to the No Mutation (46.8%) and Unknown (68.2%) groups (proportion test: *χ^2^*(2)=13.1, *p*=0.001).

**Figure 2.**
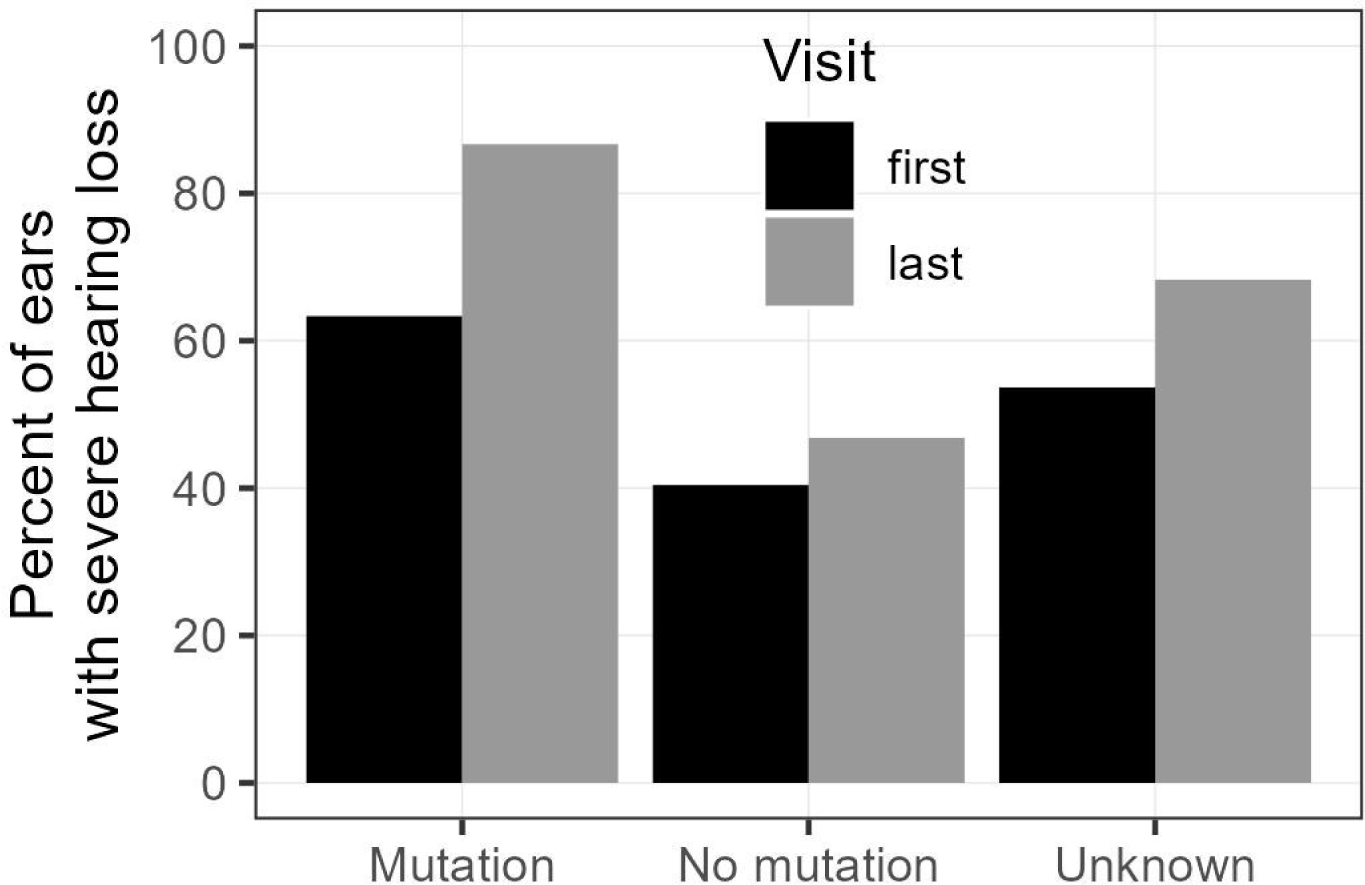
Percentage of ears with severe hearing loss (HL) at first and last visit according to SLC26A4 status

### Longitudinal analysis of hearing loss across frequencies shows a more rapid progression to severe hearing loss in the mutation group

After establishing that there was a change in the degree of hearing loss and the number of children who have severe hearing loss, our third aim sought to characterize the timing of this hearing progression. **Figure 3** shows the hearing thresholds for each ear and frequency plotted across test time in years since first visit for each individual child. As EVA can be unilateral, ears without EVA for some patients are also shown for comparison by light grey lines, but there were not enough unilateral cases to be included in the statistical analysis. The data from only ears with EVA were analyzed using a linear mixed effect model with frequency and time as fixed effects and random effects of child and type of testing. The default condition of the regression was 500 Hz for the mutation group. The model formula and results are given in **Table 2**.

**Figure 3.**
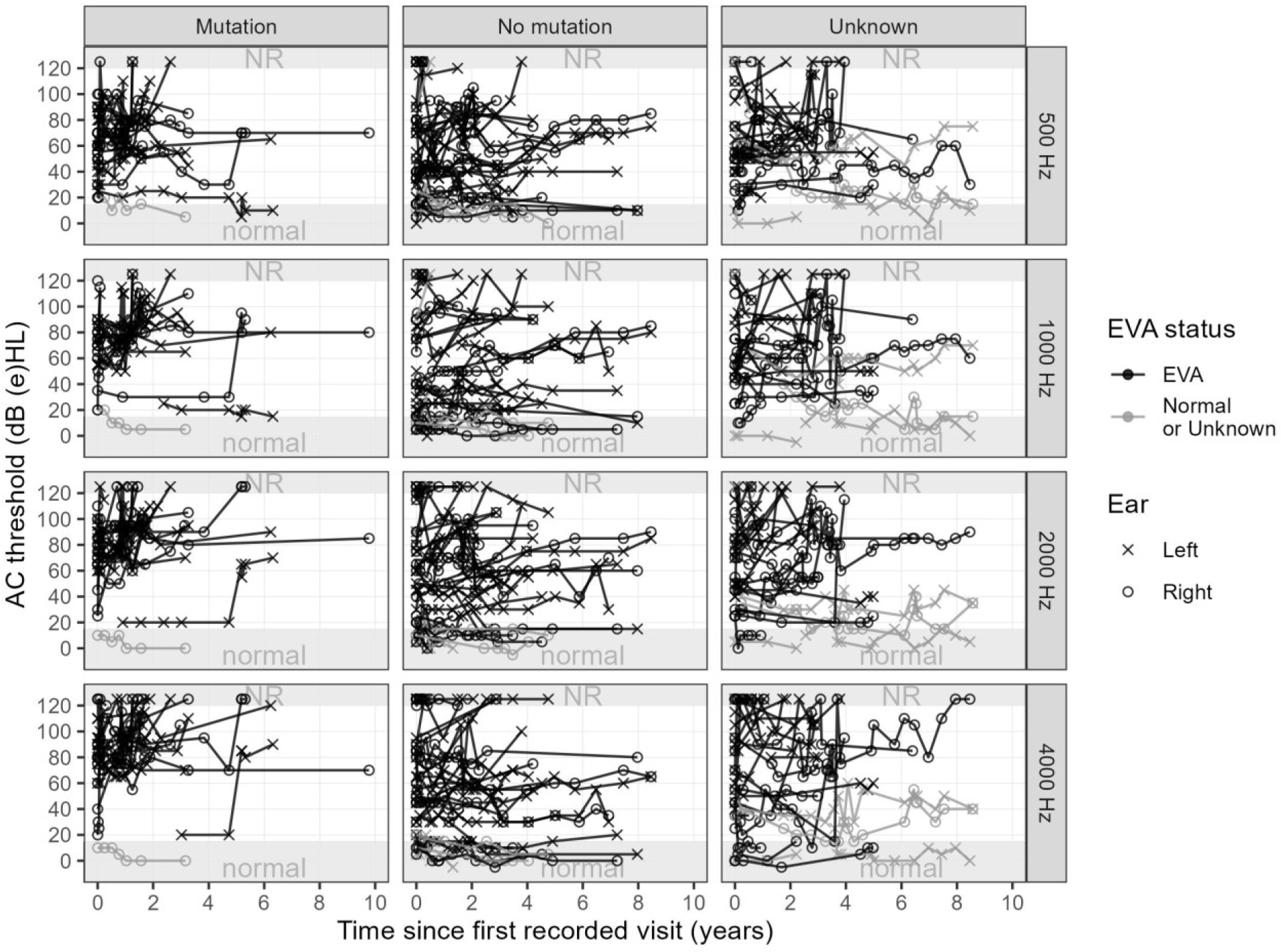
Longitudinal analysis of hearing thresholds across frequencies and time since first visit, according to SLC26A4 status. NR = No Response, Individual lines = ear for each individual child

**Table 2.** Linear Mixed Effects Model of hearing thresholds across frequency and time. Formula: threshold in dB HL ∼ time_yrs * group * frequency + (ear | child) + (1 | type of hearing test)

|  | Estimate | Standard Error (SE) | Degrees of Freedom (df) | t-Value | p-Value |
| --- | --- | --- | --- | --- | --- |
| Intercept (default: mutation group, 500 Hz) | 69.8 | 10.6 | 45.0 | 6.6 | < 0.001*** |
| Time_yrs | 1.0 | 0.9 | 2069.7 | 1.0 | 0.298 |
| No Mutation Group | -8.9 | 12.4 | 49.3 | -0.7 | 0.474 |
| Unknown Group | 5.0 | 12.8 | 50.0 | 0.4 | 0.695 |
| Frequency 1000 Hz | 7.7 | 2.7 | 2046.2 | 2.8 | 0.005** |
| Frequency 2000 Hz | 9.6 | 2.6 | 2046.1 | 3.6 | < 0.001*** |
| Frequency 4000 Hz | 12.9 | 2.6 | 2044.6 | 4.9 | < 0.001*** |
| Time_yrs: No Mutation | 0.8 | 1.2 | 2057.0 | 0.7 | 0.491 |
| Time_yrs: Unknown | 1.1 | 1.2 | 2059.6 | 0.9 | 0.329 |
| Time_yrs: Frequency 1000 Hz | 0.9 | 1.3 | 2044.4 | 0.7 | 0.504 |
| Time_yrs: Frequency 2000 Hz | 4.6 | 1.3 | 2046.6 | 3.5 | < 0.001*** |
| Time_yrs: Frequency 4000 Hz | 4.9 | 1.3 | 2043.8 | 3.9 | < 0.001*** |
| No Mutation: Frequency 1000 Hz | -7.4 | 3.6 | 2046.8 | -2.1 | 0.039* |
| Unknown: Frequency 1000 Hz | -3.5 | 3.9 | 2045.5 | -0.9 | 0.360 |
| No Mutation: Frequency 2000 Hz | -1.6 | 3.5 | 2045.4 | -0.5 | 0.651 |
| Unknown: Frequency 2000 Hz | -3.5 | 3.7 | 2045.1 | -0.9 | 0.342 |
| No Mutation: Frequency 4000 Hz | -2.1 | 3.5 | 2045.6 | -0.6 | 0.552 |
| Unknown: Frequency 4000 Hz | -6.2 | 3.7 | 2046.8 | -1.7 | 0.095 |
| Time_yrs: No Mutation: Frequency 1000 Hz | -0.8 | 1.5 | 2044.4 | -0.5 | 0.597 |
| Time_yrs: Unknown: Frequency 1000 Hz | -1.0 | 1.5 | 2044.3 | -0.7 | 0.503 |
| Time_yrs: No Mutation: Frequency 2000 Hz | -5.4 | 1.5 | 2045.9 | -3.5 | < 0.001*** |
| Time_yrs: Unknown: Frequency 2000 Hz | -5.5 | 1.5 | 2045.9 | -3.7 | < 0.001*** |
| Time_yrs: No Mutation: Frequency 4000 Hz | -8.4 | 1.5 | 2043.8 | -5.6 | < 0.001*** |
| Time_yrs: Unknown: Frequency 4000 Hz | -4.9 | 1.5 | 2044.5 | -3.3 | < 0.001*** |

Significance codes: \*\*\*P-value<0.001, \*\*P-value<0.01, P-value<0.01

As shown in **Figure 3** and **Table 2**, the Mutation group not only exhibited a more severe onset of hearing loss (model intercept ∼70 dB HL; consistent with **Figure 1A**), but their hearing loss also progressed more rapidly than the other groups, which, in contrast, had more variability in the initial degree of hearing loss and significantly shallower slopes (change in slopes, *p*<0.05). The slope for the mutation group was significantly steeper at 2000 Hz (5.6 dB/year) and 4000 Hz (6 dB/year) compared to 500 Hz (1 dB/year), suggesting faster progression for the higher frequencies. This indicates that, along with greater initial hearing thresholds (change in intercepts, *p*<0.05; 80-83 dB HL for 2-4 kHz respectively), the higher frequencies for the Mutation group reached profound hearing loss levels within 2 years of the first visit. This is illustrated by the individual lines quickly reaching the profound or “No Response (NR)” sections of the graphs in less time (**Figure 3**). These results are consistent with the increased number of children in the Mutation group who went on to receive a cochlear implant (**Table 1**). In contrast, the hearing thresholds showed little change over time in the No Mutation and Unknown groups (i.e., the threshold lines were more horizontal and the slopes were significantly smaller: ∼1 dB/year), and in some children, their thresholds showed little change even after 8 years.

## Discussion

This study investigated the changes in severity and timing of hearing loss progression in children with EVA. Of the children tested for a mutation in the *SLC26A4* gene, 38% had at least one mutation. In comparison to the group without a mutation, children with a mutation had an initially more severe hearing loss, a greater change in hearing thresholds, and a more rapid change in hearing thresholds. Consequently, more children with EVA and a mutation of the *SLC26A4* gene received a cochlear implant to support their hearing and language development. The group with unknown genetic status for this gene was highly variable and not statistically distinct from either mutation group, likely reflecting a group that contains children who both have or do not have a mutation.

The highly variable nature of hearing loss progression in patients with EVA has made rehabilitation particularly difficult (Ruthberg et al. 2022). However, our results suggest that the clinical course of EVA may be more predictable if the *SLC26A4* mutation status is known.

Specifically, children with a mutation in the *SLC26A4* gene have a more severe onset, and rapid progression of hearing loss. For this cohort, many children with a mutation progressed to a profound hearing loss within 2-3 years of their first visit and went on to receive a cochlear implant (87%). Therefore, if a child is diagnosed with EVA, clinicians may consider encouraging genetic testing of the *SLC26A4* gene when feasible to aid in the prognosis and management plan. The results of this testing would provide significant clinical value because the child’s genetic status: 1) may better inform prognostic counseling, especially when there is an identified mutation; 2) likely changes the course of hearing progression, and 3) may suggest a faster path to cochlear implantation. More specifically, our results show that genetic testing may better facilitate prognostic counseling to pediatric patients and their families – namely, if they test positive for an *SLC26A4* mutation, clinicians may recommend more frequent audiologic visits to track progression and better prepare the patient and their families for the likelihood of rapid and severe hearing loss progression.

Our results also reveal that genetic testing for *SLC26A4* mutations may suggest a sooner path to cochlear implantation to support spoken speech and language development. In our cohort, 13 of 15 children (87%) in the Mutation group received a cochlear implant by their last recorded visit (last visit median age 5.35 years old), whereas the No Mutation group had 8 out of 24 children (33%) with a cochlear implant by their last visit (last visit median age 6.8 years old. In other words, before a child with a mutation enters kindergarten, they may have a hearing loss that has progressed sufficiently to warrant conversations and referrals for implantation, but not as many would enter these conversations without a known mutation. Our results are consistent with reports of EVA and monitoring for cochlear implant candidacy in a large cohort study recently conducted in the United Kingdom (Saeed et al. 2022). Their study found that EVA patients often qualified as cochlear implant candidates in at least one ear by the age of three years. Similarly, Hodge et al. (2021) found that about 60% of children with EVA met criteria for cochlear implantation at their first audiological visit and up to 75% met criteria before 12 years old, therefore compelling clinicians to counsel parents about CI candidacy at diagnosis. Our results suggested that the hearing thresholds of children with an *SLC26A4* mutation reached profound levels within two years from their first visit to the clinic. Taken together, these findings demonstrate an *even stronger impetus* to consider a sooner path to cochlear implantation among patients with EVA who have an *SLC26A4* mutation. Clinicians may monitor children with EVA and a known *SLC26A4* mutation more closely, as their hearing loss progression will likely occur faster, and counsel the families about the potentially shorter path to cochlear implantation.

Whereas positive genetic testing results provide more specific information about hearing loss progression, a negative *SLC26A4* result also informs counseling. Our results suggest that children who do not have a *SLC26A4* mutation are less likely to have hearing thresholds that progress rapidly. Knowing this, clinicians may alter their suggestions for audiological monitoring, such as scheduling fewer follow-up appointments. Furthermore, no mutation results would also help ease any concerns on behalf of the patient and their family regarding potential rapid and severe hearing progression, as our results show that is not the case for most children (**Figure 1B**, **Figure 3** middle panel).

Characterizing the severity and timing of hearing progression brings us closer to elucidating the vast and variable puzzle of an EVA diagnosis. The results suggest that genetic testing results for an *SLC26A4* mutation are incredibly valuable and can greatly benefit clinicians in knowing how best to counsel and monitor patients. But they can also strongly benefit patients and their families in knowing what to expect – both in terms of functional outcomes and in terms of audiological follow-up timelines.

There are some limitations to this study. First, as a retrospective review, the analysis is limited to patients with sufficient and available follow-up data, and the self-selection of families who chose to participate in the database. Second, data from our cohort of 62 patients may not capture all the nuances of the larger clinical population. Thirdly, as is typical with pediatric data, not every child had every frequency threshold at each clinic visit. Despite this limitation, the data reflects what clinicians face in clinics. In the future, it would be worthwhile to conduct more research on the effect of an *SLC26A4* mutation among children with EVA in a prospective longitudinal study. Also, it would be informative to look more into genetic testing of other genes that are associated with EVA to better understand their contributions and potential combined effects on auditory outcomes.

## Conclusion

In support of our original hypothesis, the data suggest that children who have EVA and an *SLC26A4* mutation have more severe hearing thresholds, more severe onset and change in hearing thresholds, and a more rapid progression of hearing loss, especially for higher frequencies, than children with EVA who do not possess the gene mutation. This longitudinal information may facilitate better prognostic counseling to pediatric patients and their families, and better facilitate the audiology follow-up appointment schedule.

## Ethical Approval

Institutional Review Board approval was obtained from the University of Minnesota and Fairview Health Systems Research (Study 16972).

## Data Availability

The data of this study are available on reasonable request from the corresponding author. The data are not publicly available due to containing information that could compromise the privacy of research participants.

## Author Contributions

This project was conceptualized by all authors. Data curation and analyses were done by M.O. and M.P. Supervision was done by M.P., M.K., and H.H. M.O. wrote the manuscript. All authors reviewed and edited the manuscript at all stages.

## Acknowledgments

This work was funded by the American Academy of Audiology (Jerry Northern Scholarship in Pediatric Audiology to M.O.) and the Minnesota Lions Foundation Research Grant (H.H.).

